# The risks and benefits associated with the self-selection of pharmacy medicines (PMEDs): A rapid systematic review

**DOI:** 10.1101/2025.07.12.25331435

**Authors:** Lauren S. Ross, Amira Guirguis, Wing Tang, Laura Wilson, Elen Jones, James Davies, Diane Ashiru-Oredope

**Author notes:** **Corresponding author:** Diane Ashiru-Oredope. **Funding statement:** This research was conducted as part of Royal Pharmaceutical Society Science & Research Team’s regular programme of work. No specific external funding was received.

## Abstract

**Background:** Pharmacies in the UK are increasingly implementing a self-selection model for Pharmacy medicines (PMEDs) in physical pharmacies and allowing their purchase from online pharmacies. This model potentially weakens the additional level of protection recommended by the Medicines and Healthcare products Regulatory Agency (MHRA) by removing the opportunity for intervention, possibly risking patient harm.

**Objective:** To assess the risks and benefits associated with a PMEDs self-selection model in pharmacy settings.

**Methods:** A systematic search was conducted across three databases (PubMed/Medline, Embase, and Cochrane Library) from 01/10/2024 to 22/10/2024. The search terms comprised Medical Subject Headings (MeSH) terms and free text with wildcard truncations. Only studies published from 01/2014 – 10/2024 and published in English were eligible for inclusion. Studies identified (were exported to Excel, where duplicates were removed . The Mixed Methods Appraisal Tool (MMAT) was used to assess the quality of all included studies.

**Results:** A total of 55 studies of the 104 initially screened were included in the review. The country most frequently reported on was Australia (10/55; 18%), followed by multi-location/global studies (9/55; 16%). The majority (30/55; 55%) of studies focussed on the views and experiences of pharmacy professionals, and approximately a quarter (13/55; 24%) focussed on patient views, experiences, or behaviours. The benefits identified from the included literature were relatively consistent, focussing on access to care, reducing pressure on health systems, and improved patient autonomy. However, the list of risks identified was far more heterogenous, covering a range of themes, including, adverse effects of medication, inappropriate use of medication, and reduced intervention opportunities, and self-diagnoses delaying required care. The risks associated with medication self-selection were more frequently discussed when compared with benefits.

**Conclusions:** Although this review identified risks and benefits associated with medication self-selection more broadly, none of the included publications solely discussed PMEDs. More research is needed to fully understand the risks and benefits of the self-selection model for this classification of medicines.

## Introduction

In the UK, there are three classes of medicinal products for humans under the Human Medicines Regulations 2012, General Sales List (GSL) medicines, Pharmacy medicines (PMEDs), and Prescription Only medicines (POMs)^1^. These classifications are designed to ensure patients can access care with minimal risk to their health, through the required involvement of healthcare professionals (HCPs) to varying degrees. Typically, POMS are used for conditions that are formally diagnosed and managed closely by HCPs, whereas PMEDs can be advertised to the public and are for short-term treatment of medical conditions that are not chronic or likely to persist^2^. PMEDS must be sold from a registered pharmacy by a pharmacist, or by a person acting under the supervision of a pharmacist^3^. As such, PMEDs that are only available pharmacies are typically stored and displayed more carefully than GSL medicines in order to reduce the risk of patient harm. The additional restrictions recommended for PMEDs, such as limited medication volumes, and the involvement of pharmacists to assess medication appropriateness, suggests that greater caution is advisable in their sale^2^.

In community pharmacy settings, PMEDs have traditionally been stored behind the pharmacy counter, to enable pharmacist oversight and involvement in their sale. However, existing legislation does not prohibit pharmacy owners from placing PMEDs within public reach, enabling a form of self-selection^4^. This model is increasingly being adopted by brick-and-mortar pharmacies and online pharmacies, where the opportunity for direct intervention is further reduced. While the General Pharmaceutical Council (GPhC) adopts an outcomes-based regulatory approach that allows flexibility in meeting professional standards, it has clarified that open access to PMEDs (termed self-selection) is not automatically compatible with these standards. Safeguards must be in place to ensure continued pharmacist supervision and the safety of patients and the public. The GPhC favours the term “facilitated self-selection” to emphasise the role of the pharmacy team in supporting the safe and appropriate supply of PMEDs. This includes implementing controls such as tills programmed to flag PMEDs, and documented risk assessments^5^.Medication errors are the most common and easily preventable cause of patient harm^6^. Therefore, ensuring an appropriate sale model is implemented that allows easy access to medication while preventing patient harm is essential. A range of arguments have been proposed for and against the self-selection of PMEDs. Those for this model suggest that it is a more consumer-focused approach that could allow patients access to a wider selection of medication to consider while still allowing pharmacists an opportunity to refuse a sale if it would not be appropriate. There is also the argument that open selection still allows patients to read the information that is provided by manufacturers, allowing them to make an informed decision.

Those against the self-selection model emphasise that the separate classification is there to protect patients, as they have been deemed by the MHRA to require an additional level of protection because they have the potential for harm, which they believe are weakened by self-selection. Recent work from the Pharmacy Defence Association (PDA) reports that over 90% of pharmacists surveyed members (n=1,323) oppose the self-selection model of PMEDs. The majority pharmacists who responded to this PDA survey expressed concerns about the availability of appropriate supervision in pharmacies, patients’ inappropriate selection of medication, and also potential disputes or violence which may result if medication that has been self-selected is questioned or the sale is refused^7^.

While the existing work reporting on the various opinions and concerns within the pharmacy profession provide great insight, there is still limited evidence exploring the real-world consequences of implementing a self-selection model in UK pharmacy settings. Evidence is needed to fully understand the potential implications of this trending change in medication display and sale processes The primary aim of this literature review is to explore the risks and benefits associated with medication self-selection across various health systems to better understand the potential implications of applying this model to PMEDS in UK pharmacies. A secondary aim was to identify what potential harms can result from specific medicines sold via self-selection, records of specific patient behaviours in relation to PMEDs, and pharmacy experiences.

## Methods

### Database searching

This rapid systematic review of evidence was registered on PROSPERO (CRD42024600283) and follows the PRISMA 2020 mixed-methods systematic review reporting guidance (Appendix 1)^8^. The search terms shown in Table 1 were searched across three databases (PubMed/Medline, Embase, and Cochrane Library) from 01/10/2024 to 22/10/2024 to identify relevant literature for inclusion. A list of Medical Subject Headings (MeSH) terms and free text with wildcard truncations were collated to ensure the desired scope of literature was captured. The terms (Table 1) were adapted to fit the requirements of each database searched.

**Table 1.** Key search terms used in rapid systematic review search. Terms and search queries were adapted as needed across the three databases: PubMed/Medline, Embase, and Cochrane Library.

| Theme | Search Terms |
| --- | --- |
| Pharmacy location & staff | (Pharmac*) OR (online pharmac*) OR (website pharmac*) OR (remote pharmac*) OR (pharmacist*) OR (Online Pharmaceutical Service*) OR (Pharmaceutical Service, Online) (internet pharmacy) OR (e-pharmacy) |
| AND |  |
| Medication | (non-prescription*) OR (OTC) OR (over counter*) OR (pharmac*only drugs) OR (pharmac* only medication) OR (P-med*) OR (P-medicines) OR (Pharmacy medicines) |
| AND |  |
| Illness | (Self-care) OR (Self-care medicines) OR (Minor ailment*) OR (Minor illness) OR (Health management) |

To ensure our results were not duplicating the systematic review published by Eikenhorst L, et al. in 2017^9^, only studies published from 01/2014 – 10/2024 and published in English were eligible for inclusion. This allowed us to build upon the existing knowledge-base and provide a contemporary overview of the risks and benefits associated with PMEDs. This review includes findings concerning non-prescription medicines due to the limited availability of evidence relating to PMEDs specifically.

### Study criteria & data extraction

Included studies were conducted in UK and high-income countries with a similar community pharmacy context to the UK including Republic of Ireland, European Union and other relevant European countries, Australia, New Zealand, Canada, and the United States of America. Other high-income countries where the model of community pharmacy is different to the UK (e.g., Japan) were excluded. The participants/population of interest were pharmacists, pharmacy professionals, pharmacy technicians, pharmacy support staff and patients in the specified countries. The exposure of interest was the self-selection of medicines in pharmacy settings.

Studies identified using the search terms (Table 1) were exported to Excel, where duplicates were removed. The remaining records were exported to Mendeley reference managing software (v2.128.0) where the first reviewers screened all titles and abstracts based on the above inclusion/exclusion criteria^10^. Those which did not meet the inclusion criteria were discarded and documented as such. A secondary, blinded reviewer screened 20% of all exported studies to ensure appropriateness of the inclusion/exclusion process.

### Quality assessment

The Mixed Methods Appraisal Tool (MMAT) was used as a checklist for concomitantly appraising and/or describing studies included in systematic mixed studies reviews (limited to original qualitative, quantitative and mixed methods studies) was used to assess the included studies (Appendix 2). Papers which could not be assessed using the MMAT, such as systematic reviews and case series, were assessed using the JBI Critical Appraisal Tools. Included commentaries, perspectives, and evaluations were not subject to quality assessment.

## Results

Narrative synthesis and analysis were used to summarise the results due to the observed heterogeneity among the selected studies. While the primary aim of this review was to determine the risk and benefits associated with the self-selection model for PMEDs, it also aimed to identify what potential harms can result from specific medicines sold via self-selection, records of specific patient behaviours in relation to PMEDs, and pharmacy experiences. As such, the heterogeneity amongst selected studies has not been investigated and the amalgamation of data into a meta-analysis was not possible.

A total of 55 distinct studies of the 104 initially screened were identified as relevant and included in the final review (Figure 1). Forty-four studies were excluded during initial screening, and one paper could not be retrieved for full screening. An additional four studies were excluded for the following reasons: No identification of risks or benefits associated with medication (three studies); Does not concern population of interest (one study).

**Figure 1.**
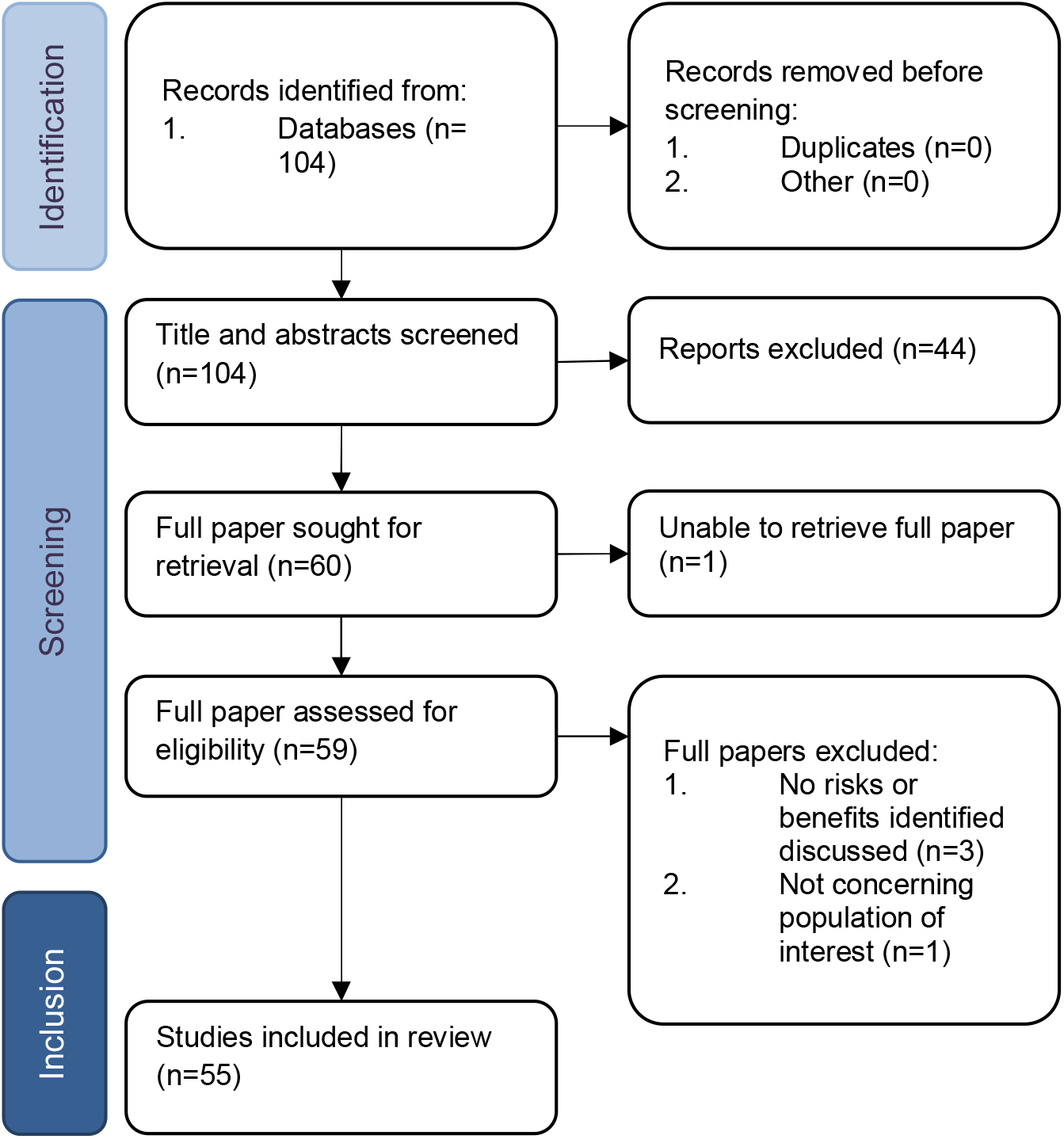
Flowchart detailing review selection process according to PRISMA guidelines

The study characteristics for the extracted papers is shown in Appendix 3.

### Study Characteristics

The 55 included studies provide insights from 24 named countries: Australia (10), United States of America (6), Poland (6), England (4), Sweden (4), Spain(3), Germany (3), Scotland (3), Canada (3), Mexico, Belgium, Croatia, Csech Republic, Denmark, Finland, France, Greece, Italy, Malta, New Zealand, Northern Ireland, Portugal, Romania, and Switzerland (1 each). 9 of the 59 studies are global studies. The majority of study participants are either of pharmacy professionals (23/55; 41%), or patients (28/55; 50%). Community pharmacies made up the majority of study settings (30/55; 54%).

### Quality Assessment

Of the included studies, 35% (19/55) were questionnaires or surveys, and a further 20% (11/55) were literature reviews of various types (including systematic reviews; 8/11), and 9% (5/55) were mystery shopper/patient simulation studies.

All eligible papers (27/55) were assessed using the MMAT (Table 2). The included studies varied in methodological rigor, with quantitative and qualitative studies scoring higher (mean MMAT scores: 70% and 72.5%, respectively) than mixed-methods studies (mean MMAT score: 53.3%).

**Table 2.**
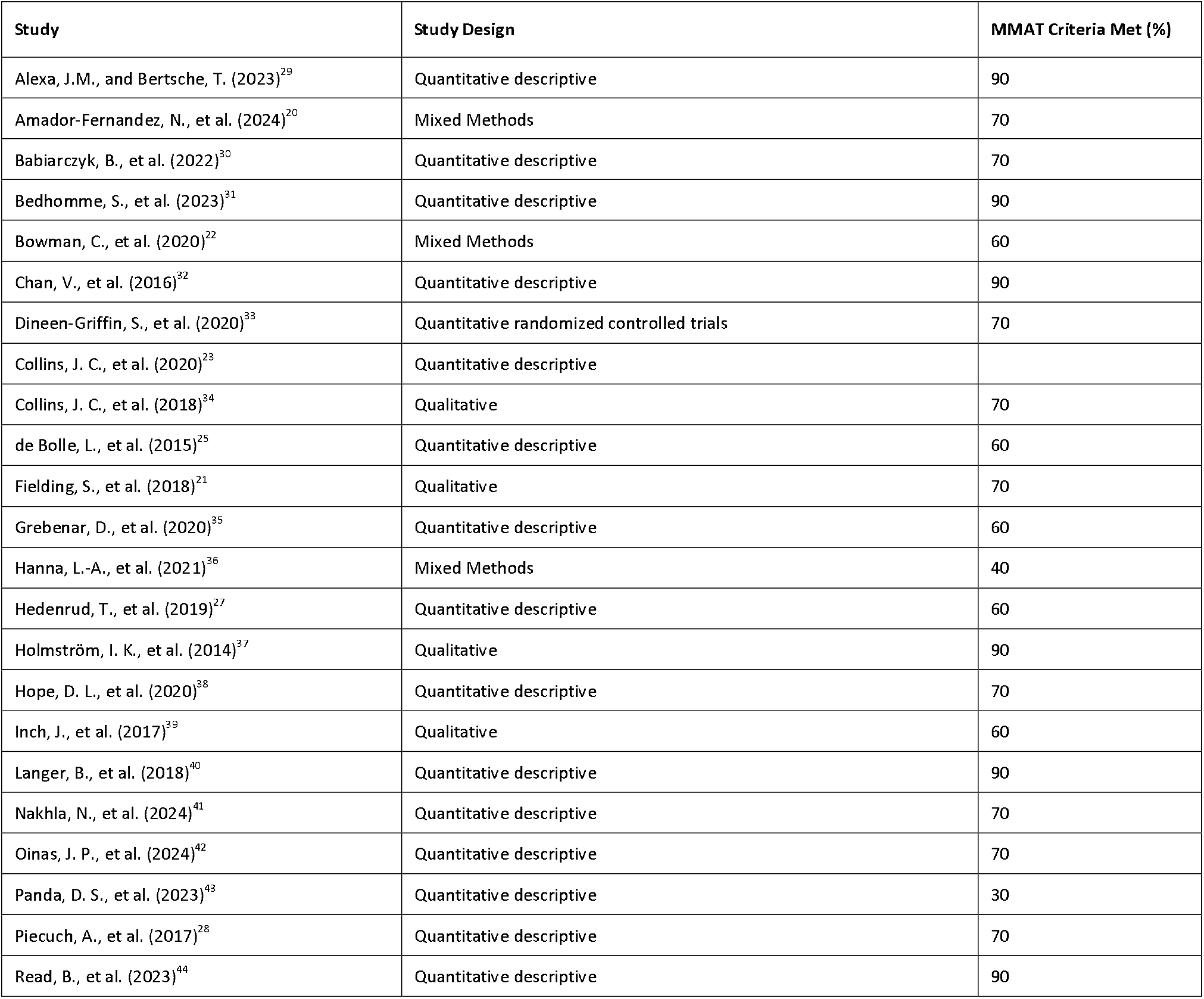
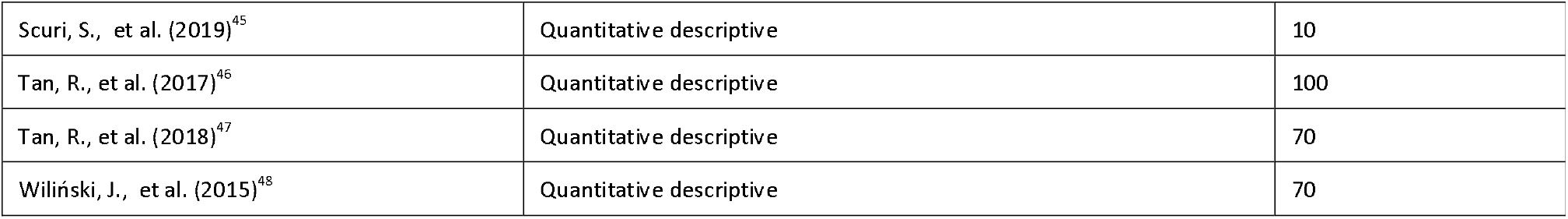
Mixed Methods Appraisal Tool (MMAT) quality assessment results for eligible papers included in review (27/55)

The included studies which weren’t assessable using MMAT (9/55), such as case series and systematic reviews, were assessed using the JBI critical appraisal tool (Table 3). The included systematic reviews varied in methodological rigour, with the proportion of criteria met ranging from 20% to 90% (mean – 61.25%).

**Table 3.** Results of the JBI Critical Appraisal Tool Checklists for systematic reviews and case series included in the review (9/54).

| Study | JBI Checklist Criteria Met (%) |
| --- | --- |
| **Systematic Reviews** |  |
| Atkins, K., et al. (2022)<sup>49</sup> | 80 |
| Jerez-Roig, J., et al. (2014)<sup>24</sup> | 80 |
| Kennedy, C. E., et al. (2019)<sup>50</sup> | 70 |
| Oleszkiewicz, P., et al. (2021)<sup>13</sup> | 20 |
| Perrot, S., et al. (2019)<sup>26</sup> | 20 |
| Seubert, L. J., et al. (2018)<sup>51</sup> | 60 |
| van Eikenhorst, L., Salema, N. E., & Anderson, C. (2017)<sup>9</sup> | 90 |
| Zheng, Y., et al. (2023)<sup>52</sup> | 70 |
| **Case Series** |  |
| Veiga, P., et al. (2015)<sup>53</sup> | 60 |

The remaining 18 studies included in this review that were not critically appraised are commentaries or quality improvement studies, which are not assessable using the MMAT or JBI critical appraisal tools. As such, the risk of bias amongst this sub-set of papers is likely to be high.

### Benefits of PMED self-selection

The recorded benefits of PMED self-selection were explored by many of the included studies, covering four core narratives. The frequency of each of the identified themes is shown in Table 4.

**Table 4.** Frequency benefit themes and associated with medication self-selection and example quotes identified from included studies.

| Benefits | n | Frequency (%) |
| --- | --- | --- |
| Convenient healthcare access | 19 | 35 |
| Reduced pressure on health systems | 7 | 13 |
| Improves self-care capability | 10 | 18 |
| Equitability of care access | 2 | 4 |

#### Convenient Healthcare Access

Twenty studies noted that the opportunity to self-select medicine increases healthcare accessibility, as patients are not restricted to prescription only following a medical consultation. This wider access to medicines allows patients to manage minor ailments independently, resulting in prompt symptom relief and no delays to care as a result of health care professional (HCP) unavailability.

*“[Medicines] are available in pharmacies, and sometimes supermarkets, enhancing convenience for patients”*^11^

#### Reduced Pressure on Health Systems

Eight studies highlighted that the availability of medicine for self-selection may help to alleviate the burden on primary healthcare providers, such as GPs. Over the counter (OTC) and PMEDs can be accessed , without the need for a primary healthcare practitioner appointment. Patients using readily available medicines to manage minor ailments can reduce waiting times for medical appointments, lower healthcare costs, and lessen the workload for doctors, supporting healthcare sustainability. During the COVID-19 pandemic, for instance, there was:

*“a sustained rise in respondents managing respiratory tract infection (RTI) symptoms with OTCs instead of consulting healthcare professionals. This trend aligns with public health messaging to reduce healthcare strain.”* ^12^

#### Improvement in Patients’ Self-Care Capabilities

Ten of the Fifty-five included studies explored how self-medication empowers patients to take control of their health by addressing common conditions autonomously, promoting self-reliance. Improved access to medications allows individuals to take control of their health and address common, mild ailments without the need to see a primary care provider. This makes healthcare more convenient, timely, and patient-driven. This can be especially important for easily manageable, chronic conditions (e.g., heartburn) which can impact day-to-day life without intervention:

*“The prevalence of self-medication leads to a decrease in the share of healthcare in the treatment of mild disorders and has a positive impact on the quality of life of patients suffering from chronic and recurrent diseases. Examples where self-treatment can be implemented include a cold or flu, digestive disorders (including heartburn), and mild to moderate pains such as headaches and muscle pains”*^*13*^

^*14*^*Equitability of Care Access*

The advancements in self-care services have improved the equitability of healthcare access by offering interactive, patient-centred resources that facilitate autonomous healthcare decisions. Resources such as general practice apps have improved the accessibility of information on minor ailments and advice on the best course of action to take without having to consult a doctor directly. This is noted as an advantage in two of the included studies and is highlighted as being particularly significant for individuals who may face barriers to traditional healthcare access or seldom-heard patients.

*“Expansion of digital self-care services improve care efficient and allows patients to take a more active role in their health management. Creates potential for more interactive, patient-centred, equitable healthcare systems*.*”*^*14*^

### Risks of PMED self-selection

Seven themes were identified relating to the risks associated with medication self-selection in the 55 included studies from this review. The frequency of each of the identified themes is shown in Table 5.

**Table 5.** Frequency of risk themes and associated with medication self-selection and example quotes identified from included studies.

| Risks | n | Frequency (%) |
| --- | --- | --- |
| Adverse Events | 18 | 33 |
| Inappropriate use of medication | 17 | 30 |
| Health literacy | 6 | 11 |
| Self-diagnoses delaying medical attention | 8 | 14 |
| HCPs' education/training on clinical evidence | 4 | 7 |
| Limited opportunity for pharmacist intervention | 3 | 5 |
| Counterfeit medications | 2 | 4 |

#### Adverse Events

Nineteen studies identified the occurrence of adverse events as a risk associated with OTC medication access, particularly for high-risk patient groups such as immunocompromised individuals, elderly patients and poly-medicated patients. If patients are poly-medicated, the use of self-selected medications may be contraindicated by existing medication, resulting in serious adverse events.

Adverse events can occur in all patient types, not just those who are high-risk. Commonly reported adverse events included gastrointestinal symptoms (nausea, vomiting) and central nervous system issues (anxiety, insomnia). These symptoms can be distressing for patients and may not be immediately associated with the self-selected medication.

#### Inappropriate Use of Medication

Seventeen studies highlighted that patients may struggle with selecting the appropriate medication for their condition. Lack of knowledge about symptoms, product indications, and the mode of action of medications can lead to misuse of medication. Bertsche, T. et al.’s study found evidence that suggests that as much as 12% of OTC products sold in Germany have a potential for misuse^15^.

#### Self-Diagnosis Delaying Medical Attention

Patients may identify symptoms and associate them with the incorrect ailment, addressing them with self-selected, readily available medication rather than seeking appropriate care. Taking a PMED or GSL medicine to address inconspicuous symptoms can mask the progression of a serious conditions, delaying necessary medical attention.

#### Health Literacy Challenges

Limited health literacy was noted as a barrier to safe self-medication in six of the fifty-five included studies. Some studies highlighted that individuals lack knowledge about potential OTC side effects or are unable to completely understand the information provided on the medication packaging. The lack of literacy and/or knowledge can result in patients taking the incorrect medication for their symptoms and/or condition, allergic reactions, contraindications with other medication, or unexpected side effects. Without effective consultation and tailored guidance on medication selection, there is a risk of harm to patients.

#### Healthcare Professional (HCP) Education and Training Gaps

Four studies raised concerns focussed on HCPs education and training. For example, one study suggested that HCPs may lack adequate knowledge of the clinical data for some OTC products, affecting their ability to counsel patients effectively. The lack of knowledge or experience with medications may limit pharmacy teams’ ability and confidence to intervene or challenge the purchase of self-selected medication.

#### Limited Opportunity for Pharmacist Intervention

A few studies (3/55) highlighted the limited opportunities for pharmacist intervention when it comes to medication self-selection. Staff availability, sale at self-checkouts, and subject matter were noted as factors which may limit intervention opportunity. One study found that certain topics may make the patient and/or pharmacist uncomfortable, leading to ineffective counselling^9^. This reliance on informational pamphlets as opposed to face-to-face discussion reduces the opportunity for pharmacists to advise on the self-selection of medications.

#### Counterfeit Medications

The rise of unregulated online pharmacies has led to concerns about the availability of counterfeit medication. This raises additional safety concerns for self-selected medication as, if purchased from un-verified sources, a patient may be unintentionally taking a dangerous or substandard product, which bears no similarity to what they ordered. This risk was mentioned in two of the fifty-five studies.

## Discussion

There is limited published literature exploring the risks and benefits associated with PMEDs specifically; however, this review has gathered evidence relating the self-selection of all OTC medication types, and patients’ relationships with medication and self-care. The included studies provide insight into how patients use easily accessible medications and discuss the risks and benefits associated with self-medication.

This review found that the benefits associated with medication self-selection were primarily focussed on individuals’ access to care. This increased access to care came with improved autonomy for patients, especially in the management of mild conditions which do not require a practitioner’s input. This aligns closely recently published Welsh strategy for health and social care, which emphasise the importance of empowering patients to be more involved in their own health and wellbeing in order to keep the wider population healthier and reduce the burden on existing health services^13^.The ease of access to medication instils a sense of autonomy in individuals, allowing them to take control of their health and make their own health decisions. Bertsche, T., et al (2023) noted this, and suggested that this is a more convenient way of managing common, transient health conditions.^16^ This study also noted that the self-selection of medication also reduces the pressures placed on healthcare professionals, in cases where their involvement is likely not required and there is minimal patient risk. The overall reduction in health systems pressure was a very common benefit, with several publications pointing to the common understanding that healthcare professionals are typically overworked. ^17–19^ The easier access to medication would, therefore, allow the limited resources to be reallocated within the healthcare system, to more critical services. However, the lack of involvement of pharmacy professionals in patient’s treatment does remove the opportunity for intervention. Although viewed as a benefit, the removal of pharmacy professionals’ input to the medication selection process could pose a risk.^9,20,21^ The exact circumstances in which the self-selection of medicine can improve patient’s care access and autonomy without negatively influencing their health outcomes requires further assessment, as this balance is not yet fully understood.

This pool of data collated in this review suggests the existence of potential harm associated with medication self-selection models. The majority of the identified risk themes (Table 5) relate to patient’s health outcomes, as a result of medication misuse or sourcing medication from illicit sources. The most prominent concerns are focussed on the occurrence of adverse events and potential contraindications, which can have significant health consequences for patients. By removing the involvement of qualified healthcare professionals from the medication selection process, patients are not able to receive advice on which medication is most appropriate for them, not only based on their symptoms, but also based on their current medications and medical history.

This was noted throughout the included studies, which primarily focussed on GSL products; however, it is likely that this would extend to PMEDS. Additionally, this is likely to have greater potential consequences due to the increased risks of harm associated with PMEDs^3,5^. Removing the provision of guidance based on these patient-specific factors may result in patient harm. Interestingly, publications, such as Bowman, C., et al. (2020) found that patients believe there is less risk associated with easily accessible medications, such as OTCs, compared to other medication classifications.^22^ This lack of perceived risk may lead to inappropriate medication purchases. The implementation of a self-selection model for PMEDs may result in these medications being viewed in a similar light, with patients not fully aware of the potential harm that could result from their inappropriate use.

As well as adverse reactions, incorrect self-diagnosis may mask symptoms of serious illnesses, delaying proper diagnosis and treatment by a healthcare professional.^17,23–25^ This has dangerous implications for the patient and can inadvertently increase the burden on healthcare systems, as undiagnosed conditions may escalate, requiring more intensive and costly medical interventions further on. In addition, the condition being masked or insufficiently treated may progress and cause significant health decline before the patient seeks appropriate care, resulting in more emergent conditions. This is of particular concern in vulnerable patients, such as those who are elderly or polymedicated.^24^

Similarly, the lack of training and availability for pharmacists and wider pharmacy teams was noted to reduce their capability to intervene in inappropriately self-selected medication sales, also risking potential harm to patients^19,26–28^. From what is presented in the identified literature, the risks can cause significant harm to patients, but can be addressed by appropriate interventions, such as improved pharmacy staff training, clarification of role responsibilities, and more clear, obvious product labelling. The included literature reports these themes as risks due to the barriers which limit the ability to address these shortfalls in staff capability or material clarity^28,29^. These results reinforce the results published by Eikenhorst, et al. (2017), who found that counselling on medication selections was inconsistent and, when provided, insufficient in addressing potential risks.^9^

The accessibility of the self-selection of medication was underscored during the COVID-19 pandemic, wherein pharmacies remained open, providing both advise and OTC medications when other healthcare services were strained^9^. For patients, this convenience translated to saved time, decreased work absenteeism, and lower prescription costs, all of which enhanced their overall quality of life. Moreover, self-selected medications often present more affordable alternatives, increasing accessibility for consumers.

For common ailments, for which standard of care is commonly understood among wider populations, access to medication allows patients to act autonomously and take control of their care promptly, removing an additional step which delays care access and burdens both the patient and health care system.

Since Eikenhorst, et al. (2017)’s publication, progress had been made in specifying the risks associated with limited pharmacist intervention and medication self-selection. However, the evidence base relating to PMEDs specifically is still extremely limited. The literature identified lacked documented incidents of harm and primarily provided acknowledgement of potential risks (e.g., adverse events, misuse) and safeguarding strategies (supervision, higher-risk exclusions). Further evidence is required to confirm the impact of a PMED self-selection model on patient outcomes, and the real-world impact the self-selection model has on pharmacy settings.

## Conclusions

This literature review has identified both benefits and risks associated with the self-selection of medications. Benefits such as increased healthcare accessibility demonstrates the potential positive impacts for patient empowerment and care equity. However, the risks, including adverse events, inappropriate use, and health literacy barriers, underscore the need for targeted education initiatives and improved training for pharmacy professionals and wider pharmacy teams to ensure the self-selection of medication poses minimal risks to patients.

The current pool of evidence collated in this review is insufficient to draw conclusions relating to the use of PMEDs, as none of the evidence explored the consequences PMED self-selection specifically. Much of the literature instead discussed general sale/over-the-counter medications. As such, our results discuss the risks and benefits of medication selection more broadly, rather than the PMED classification of interest. As such, it is not possible for this review to conclude whether the benefits outweigh the risks of a self-selection model for PMEDs. Moreover, the heterogeneity of the study designs made comparison between publications challenging. Additionally, the sample of studies collated was limited to those available on the PubMed/Medline, Embase, and Cochrane Library databases, and published in England. This may have resulted in the inadvertent exclusion of relevant publications. Finally, due to the variation in study design within our included publications, multiple quality appraisal tools had to be used.

More targeted research is required to better understand the risks and benefits associated with the various sale models pharmacies can implement for PMEDs, focussing primarily on patient safety and health improvement, rather than economic factors. More information is needed to determine which sale model is best suited to provide equitable, safe access to medicines. Such research will also provide essential information that can guide how pharmacy professionals should be educated and supported ahead any changes made to sale models in pharmacy settings.

## Data Availability

All data produced in the present work are contained in the manuscript

## Acknowledgements

Professor Parastou Donyai is acknowledged for her contributions to previous work, from which this systematic review builds.

